# The factors associated with HIV Pre-Exposure Prophylaxis awareness and use among women aged 40-65 years in SE England: A protocol for the analysis of data from a cross-sectional population survey

**DOI:** 10.1101/2025.09.27.25336800

**Authors:** Kiersten Simmons, Carrie Llewellyn, Stephen Bremner, Nigel Sherriff, Kate Gilchrist, Massimo Mirandola, Jorg Huber, Richard de Visser, Kathleen Galvin, Louise Knight, Catherine Aicken, Alexandra Sawyer, Collins Iwuji

## Abstract

**Background:** New HIV diagnoses are increasing among women aged 40-65 years in England. HIV Pre-Exposure Prophylaxis (PrEP), the use of antiretrovirals for HIV prevention, is an effective chemoprophylactic strategy, as part of a comprehensive prevention package, for HIV. Granular data on knowledge about, and access to HIV PrEP for women aged 40-65 years, is lacking. This project will be a secondary analysis of a survey of adults in Brighton and Hove, England. It aims to investigate the sociodemographic and sexual behaviour factors that are associated with HIV PrEP awareness and HIV PrEP use in women aged 40-65 years in Brighton and Hove.

**Methods:** ‘Health Counts 2024’, a cross-sectional, self-completion health-related survey was conducted in Brighton and Hove in March 2024. The primary outcomes will be HIV PrEP awareness and HIV PrEP use among women aged 40-65 years in Brighton and Hove and will be analysed using descriptive statistics appropriate to distribution. The secondary outcomes will be to determine whether there are associations between sexual behaviours and/or sociodemographic factors and HIV PrEP awareness and HIV PrEP use for women aged 40-65 years in Brighton and Hove. These will be analysed using descriptive statistics and multifactorial logistic regression models from which odds ratios and 95% confidence intervals will be reported.

**Ethics and dissemination:** Ethical approval for the ‘Health Counts’ survey was obtained from the NHS Health Research Authority (HRA: Ref: 23/LO/0825) and the London – Bromley Research Ethics Committee (Ref: 23/LO/0825). Ethical approval for the community enrolment and supported completion pathway was granted by the University of Brighton Cross-School Research Ethics Committee C (2023-12553). Findings from this analysis will be disseminated to stakeholders via a scheduled public engagement event (‘Valuing the voices of women in coastal communities’), presentations at conferences and in peer-reviewed journals.

**Strengths and limitations of this analysis:**

- We will be the first to examine HIV PrEP awareness and HIV PrEP use, and sociodemographic and sexual behavioural correlates among women aged 40-65 years in the UK.
- ‘Health Counts 2024’ was extensively developed, piloted, and tested to ensure accessibility and therefore representativeness of results.
- Lack of HIV PrEP awareness and HIV PreP use among women may limit statistical analysis.
- Cultural sensitivity and stigma around sexual health behaviour may limit response rates on relevant questions and therefore may limit statistical analysis.
- Generalisability of the findings may be limited due to known regional differences in population awareness of HIV PrEP.

## INTRODUCTION

Globally, women and girls account for a substantial portion of the HIV epidemic [1, 2]. In England, despite national improvements in reducing HIV transmission to meet the target to eliminate HIV transmission by 2030, progress for women has been disproportionately slow [3]. The rate of new sexually transmitted infection (STI) diagnoses, and the number of new HIV diagnoses in women in the United Kingdom (UK) is increasing [4].

HIV Pre-Exposure Prophylaxis (PrEP), the use of antiretrovirals for HIV prevention, recommended for individuals at substantial risk of HIV infection [5], has demonstrated efficacy in preventing HIV transmission in clinical settings worldwide as part of comprehensive prevention methods [6–8]. Women account for 27% of all new HIV diagnoses in England, but only represent 2% of current HIV PrEP users [9]. Early HIV PrEP trials complicated health promotion efforts for women as they did not demonstrate significant reductions in HIV seroconversion among women [6]. The failure of these trials was largely attributed to poor adherence to HIV PrEP for many sociocultural reasons including perceived stigma, lack of social support, and a low perception of HIV risk [10]. Studies have also demonstrated the need for higher levels of HIV PrEP adherence by women (compared to men) to achieve adequate drug levels in the female genital tract tissues [10]. Although women have repeatedly been under-represented in work in this field, and most HIV PrEP services are tailored to men who have sex with men [11], multiple subsequent trials have demonstrated the effectiveness of HIV PrEP in preventing HIV acquisition among women [12]. There are subpopulations of women who could potentially benefit from HIV PrEP, but there is insufficient evidence regarding the enablers and barriers to access of the various formulations of HIV PrEP that are currently available for women [9, 13, 14].

Midlife (40-65 years) is a time of important physiological and societal changes for women, and a stage at which the trajectory of health and wellbeing into older age can be positively influenced [15]. However, midlife women are a particularly under-researched population in terms of access to sexual health and sexual wellbeing (SHSW) services, and a granular understanding of the SHSW needs of midlife women in England is lacking [16]. HIV PrEP is currently only publicly available in England from SHSW services [17]. There are significant barriers to midlife women accessing SHSW services, which are compounded by intersecting disadvantages [11, 18]. These include: intrapersonal barriers such as poor knowledge and awareness about SHSW and SHSW services; interpersonal and cultural barriers including gendered and ageist norms; and healthcare system barriers such as insufficient knowledge, skills, and attitudes of Health Care Professionals, and lack of prioritisation of the needs of midlife women within service designs. Although the UK government, supported by the UK’s most recent HIV PrEP guidelines [7], has recommended a series of pilot services to provide HIV PrEP in a variety of settings, including online, pharmacies, and General Practices, it is unclear how these would successfully be incorporated to improve equity of access for midlife women [3, 17]. Midlife women who live in underserved regions and who are from underserved groups have intersecting barriers to accessing SHSW services [19]. An understanding of whether sexual health behavioural factors and/or sociodemographic factors are associated with awareness and use of HIV PrEP among midlife women will enable better tailoring of educational and public health interventions.

## METHODS

### Primary objective

To determine HIV PrEP awareness and HIV PrEP use among a population of women aged 40-65 years residing in Brighton and Hove or registered with a GP within the city council boundaries.

### Secondary objectives

To describe the sexual health behaviour of a population of women aged 40-65 years in Brighton and Hove.

To explore the extent to which sexual behaviour factors and sociodemographic factors are associated with HIV PrEP awareness and HIV PrEP use among a population of women aged 40-65 years in Brighton and Hove.

### Research questions

Q1a: What proportion of women aged 40-65 years in this sample are aware of HIV PrEP? Q1b: What proportion of women aged 40-65 years in this sample use HIV PrEP?

Q2: What are the sexual health behaviours of women aged 40-65 years in this sample?

Q3a: Which sociodemographic and/or sexual behaviour factors are associated with HIV PrEP awareness in this sample?

Q3b: Which sociodemographic factors and/or sexual behaviour factors are associated with HIV PrEP use in this sample?

### Study design

Secondary analyses of data from participants in the ‘Health Counts 2024’ study [20] which was led by the University of Brighton in collaboration with Brighton & Sussex Medical School, Brighton & Hove City Council Public Health team, NHS Sussex, Healthwatch Brighton & Hove and, the Brighton & Hove Federation of Primary Care - as well as all GP practices in the city, and the community and voluntary sector. The aim of the study was to provide comprehensive public health-relevant data from an adult population in a defined geographic area of England [20]. The study design, data collection methods, and the inclusion/exclusion criteria for the ‘Health Counts 2024’ study has been described previously [20] The main procedures applicable to the current analysis are briefly outlined below. ‘Health Counts 2024’ was a cross-sectional, self-completion (predominantly online) health and lifestyle population survey of adults. The final survey, following extensive rounds of consultation and testing, comprised 102 questions structured around 13 public health issues in the UK [20]. The main method of recruitment had a mobile-first design approach, in the form of information and the survey link being sent by text message from General Practices (GPs) to eligible registered patients. Wider marketing (promotion through partner organisations, social media campaign, statistical monitoring of survey completion with adjusted promotion) and targeted outreach activities, were also conducted to raise awareness of the study in the general population and to maximise the potential of those often underrepresented in research to participate, including those less likely to be registered with a GP practice. This protocol describes a secondary analysis of this dataset to investigate the sexual health behaviour of a population of women aged 40-65 years and to assess whether HIV PrEP awareness and use among women aged 40-65 years in Brighton and Hove are associated with sexual health behaviours and/or sociodemographic factors.

### Study setting

‘Health Counts 2024’ was conducted in the city of Brighton and Hove, a coastal city in Southeast England, United Kingdom. There are significant inequalities in health, wellbeing and deprivation across different neighbourhoods of the city [20]. There was no sample size calculation for this convenience survey. As of the Census 2021, the population of Brighton and Hove was 277,200 (141,600 females, 135,600 males). The overall response rate to the survey was estimated to be 10.84% [22]. This was based on the population of adults registered at GPs in Brighton and Hove, with adjustment for the unknown proportion of these who were likely to have moved away or died, and the adult population size, from population projections based on Census 2021 data. 26,014 cases were included for analyses in the final dataset, of which 3361 are eligible for inclusion in these analyses.

### Participants

The study population for Health Counts 2024 was defined as anyone aged 18 years and over residing either temporarily or permanently, or registered with a General Practitioner (GP), within the Brighton and Hove city council boundary. The data of the participants who self-identified as female aged 40-65 years (inclusive) will be extracted from this survey. The inclusion criteria for these analyses are: self-identifying females; aged 40-65 years (inclusive); currently living in Brighton and Hove permanently or temporarily (e.g. students, Travellers, refugees, and asylum seekers) or living elsewhere but registered at a GP located within the City of Brighton and Hove. The exclusion criteria for these analyses are: do not identify as female; aged less than 40 years or over 65 years; responses indicate residence outside of the City of Brighton and Hove and not registered with a GP in the City of Brighton and Hove.

### Patient and public involvement

A Patient and Public Involvement and Engagement Participation (PPIEP) group consisting of five diverse midlife women for an umbrella project, ‘Sexual health and sexual wellbeing services for midlife women’ was consulted about the specific research question of exploring the sociodemographic and sexual health behaviour factors associated with HIV PrEP awareness and HIV PrEP use. The PPIEP group has also agreed to assist with the dissemination of the results. The initial survey had extensive PPIEP involvement, including pre-testing and piloting of the survey [20].

### Data Collection

‘Health Counts 2024’ opened at 07:00 on Monday 18th March 2024 and ran for six weeks until the 28th of April 2024 at 20:00. The survey was hosted on a University of Brighton landing page www.brighton.ac.uk/research/health-counts-2024.aspx [20]. The survey data will be anonymised and then extracted by the ‘Health Counts 2024’ administrator and shared, on application, via SPSS [21] with the research team. SPSS will be used to analyse the data.

### Independent (exposure) variables

*Sociodemographic measures* (Table 1) will be: age; disability; neurodiversity; level of education; current gender identity; sexual orientation; ethnicity; religion; refugee/asylum status; Index of Multiple Deprivation quintile of area of residence; partnership status. For these analyses, these variables will be categorised and displayed as per Table 1.

**Table 1:**
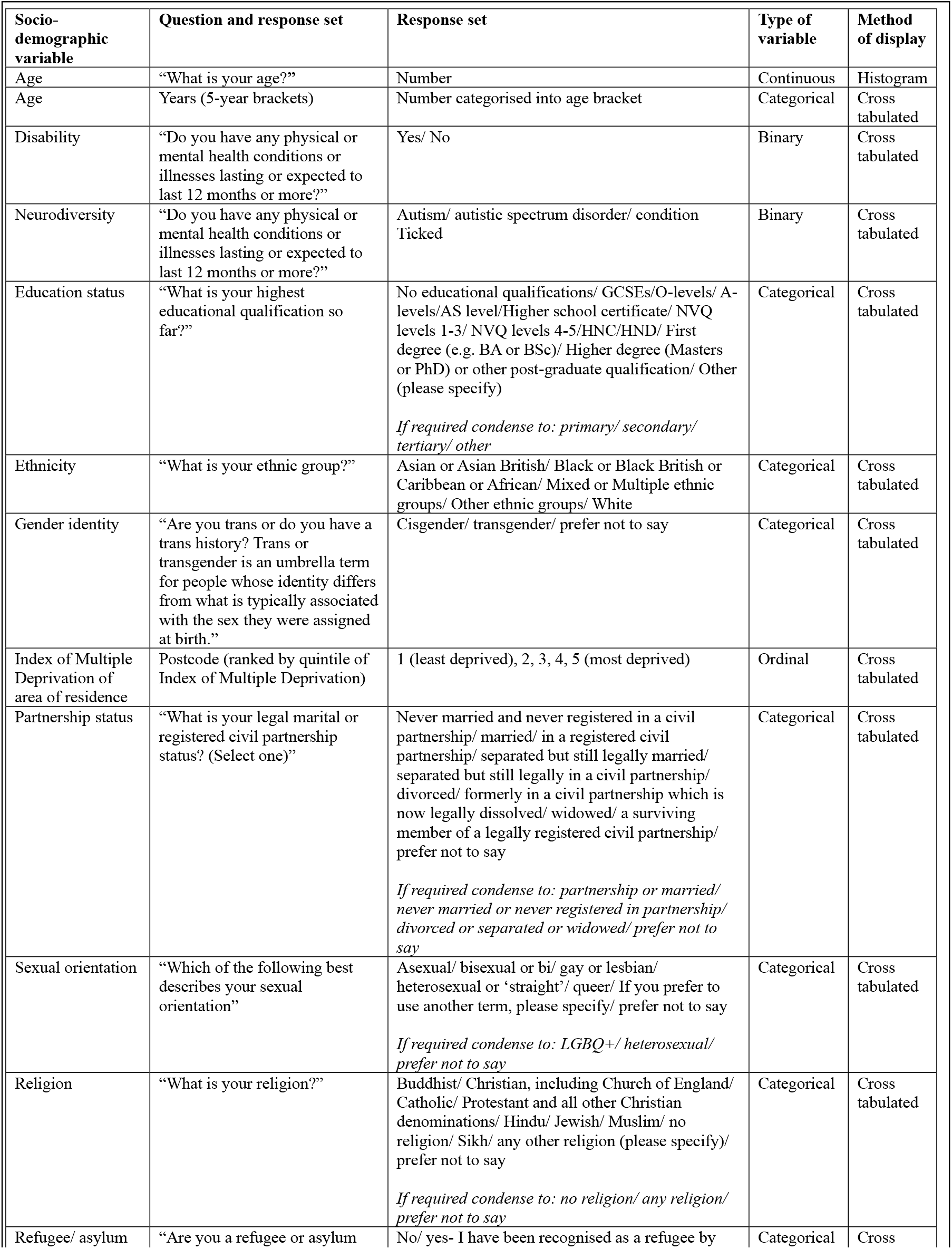
Sociodemographic variables.

*Sexual behaviour measures* (Table 2) will be: number of partners in the last 12 months; number and type of new partners in the last 12 months; condom use; HIV testing ever; HIV testing recency. For these analyses, these variables will be categorised and displayed as per Table 2.

**Table 2:** Sexual health behaviour variables.

| Sexual behaviour variable | Question and response set | Response set | Type of variable | Method of display |
| --- | --- | --- | --- | --- |
| Number of partners in the last year | “In the last 12 months, how many people have you had sex with? Include people you have had any kind of sex with (vaginal, oral, anal).” | Numerical value | Discrete numerical variable | Bar chart |
| Number and type of new partners in last year | “How many of these partners were new partners, who you had sex with for the first time in the last year?” | Men/ women/ transgender/ non-binary (numerical value)<br>*If required will condense to one numerical value (for all gender identities)* | Discrete numerical variable | Bar chart |
| Condom use | “When you have sex with a new partner for the first time, do you use a condom?” | Every time/ sometimes/ never/ not applicable/ prefer not to say | Categorical | Cross tabulated |
| HIV testing ever. | “Have you ever had a test for HIV?” | Yes/ no/ not sure/ prefer not to say | Categorical | Cross tabulated |
| HIV testing recency | “When was your most recent HIV test?” | Less than three months/ three months - twelve months/ more than twelve months <=sixty months/ more than sixty months/ prefer not to say | Discrete numerical data | Bar chart |
| HIV PrEP awareness | “PrEP (pre-exposure prophylaxis) can reduce your chance of getting HIV from sex or injection drug use. When taken as prescribed, PrEP is highly effective for preventing HIV. Have you heard of PrEP?” | Yes/ no/ I don't know | Categorical | Cross tabulated |

### Dependent (outcome) variable

We will use the categorical response to the question about HIV PrEP awareness as our response variable. 1. “PrEP (pre-exposure prophylaxis) can reduce your chance of getting HIV from sex or injection drug use. When taken as prescribed, PrEP is highly effective for preventing HIV. Have you heard of PrEP?” The response set included yes/no/I don’t know. ‘I don’t know’ responses will either be excluded, or the response set will be condensed into yes/no (including I don’t know). 2. “Have you used PrEP?” The response set included no/yes, currently/yes, in the past/prefer not to say. Due to the small number of participants in our population who are currently using PrEP, and who have ever used PrEP, it is likely that these two variables will be collapsed into one ever-use of PrEP variable i.e. no/yes. ‘Prefer not to say’ responses for each variable will be analysed with consideration to the survey and question design. For example, if the survey directs participants to skip questions that are deemed not relevant to their circumstances, ‘prefer not to say’ responses will be considered in relation to the likely response to the previous question. For other variables, it may be deemed appropriate to treat ‘prefer not to say’ responses as missing data or to conduct a sensitivity analysis.

### Data Analyses

We will use descriptive statistics to determine how the sample of women aged 40-65 years (age, disability, ethnicity, partnership status, religion, refugee/asylum seeker status, gender identity, sexual orientation, Index of Multiple Deprivation of area of residence, determined by postcode) compares with data that is publicly available for women aged 40-65 years in England (Office for National Statistics [22]). We will use descriptive statistics to answer our first research questions:

Q1a: What proportion of women aged 40-65 years in this sample are aware of HIV PrEP?

Q1b: What proportion of women aged 40-65 years in this sample use HIV PrEP?

Q2: What are the sexual health behaviours of women aged 40-65 years in this sample?

To explore the relationships between sociodemographic factors and sexual behaviours and HIV PrEP awareness and HIV PrEP use, we will consider the following research questions using logistic regression models.

Q3a: Which sociodemographic and/or sexual behaviour factors are associated with HIV PrEP awareness in this sample?

Q3b: Which sociodemographic factors and/or sexual behaviour factors are associated with HIV PrEP use in this sample?

### Statistical procedure

The data will be analysed using IBM SPSS [21].

### Descriptive statistics

1. Percentages will be used to summarise categorical variables. *Sociodemographic description of sample* (age in 5-year categories, disability, neurodiversity, ethnicity, partnership status, religion, refugee/asylum seeker status, gender identity, sexual orientation, Index of Multiple Deprivation quintile of area of residence). *Sexual behaviour description of sample* (number of partners in the last year, number of new partners and new partner type in the last year, condom use (new partners), and HIV testing (ever, recency)) HIV PrEP awareness, HIV PrEP use (current/ever)).
2. Median and interquartile range will be used to summarise continuous variables with a skewed distribution whilst mean and standard deviation will be used to summarise continuous variables that are normally distributed.
3. Number and percentage of missing values per variable.

### Assessing associations with HIV PrEP outcomes

#### Which sexual health behaviour factors and sociodemographic factors are associated with HIV PrEP awareness and HIV PrEP use?

The tests described here are aimed at finding any predictive associations and identifying likely explanatory variables. Candidate independent factor selection is based on prior literature, clinical knowledge and experience, and expert consultation (6, 19-21). Independent factors will be sociodemographic factors (age, disability, ethnicity, partnership status, religion, gender identity, sexual orientation, education, refugee/asylum seeker status, Index of Multiple Deprivation quintile of area of residence) and sexual behaviour factors (partner numbers and type, condom use, HIV testing ever and recency). For HIV PrEP use, HIV PrEP awareness will also be a covariate. To assess whether sociodemographic factors and/or sexual behaviours are related to the dependent variables of interest (HIV PrEP awareness and HIV PrEP use), we will conduct crosstabulations and descriptive univariable analyses for categorical data. Bar charts with descriptive univariable analyses will be conducted for discrete numerical data. We will conduct multivariable binary logistic regression models [23]. Factors that are significant in the univariable models will be included in the final multivariable logistic regression models. Significance will be assessed using a p-value <0.2, but also, with the recognition that p-values will only provide tentative evidence about our data [24], a priori expectations will be used. Existing evidence has suggested that age [9, 25–27], ethnicity [9, 13, 27–30], and educational level [27, 31–33] should be included in the final multivariable model regardless of statistical significance. We will investigate the magnitude and direction of associations of the sociodemographic factors and the sexual behaviours. We will report the estimated odds ratios and 95% confidence intervals from each model. We will assess the adequacy of the logistic regression models by examining the model chi-square (Wald) statistic [34]. Attrition, non-response and missing data will be examined and dealt with at the appropriate level of analyses. Findings will be reported according to the Strengthening the Reporting of Observational Studies in Epidemiology (STROBE) [35] guidelines.

### Discussion/Impact

HIV PrEP is an important intervention in the drive to eliminate HIV by 2030. There is increasing global, national, and regional recognition of the necessity to address poor and inequitable access to sexual health and sexual wellbeing care for midlife women, including access to HIV PrEP. The aim of determining the factors that are associated with awareness and use of HIV PrEP among midlife women is to support and improve appropriate public health targeting of interventions and education for those at most risk in this population.

## Ethics and dissemination

Ethical approval was obtained from the NHS Health Research Authority (HRA: Ref: 23/LO/0825) and the London – Bromley Research Ethics Committee (Ref: 23/LO/0825) for promotion of the study via SMS in general practices. Ethical approval for wider promotion of the study in the population, the community recruitment and supported completion pathway was granted by the University of Brighton Cross-School Research Ethics Committee C (CREC-C; Ref: 2023-12553-Aicken Health Counts 2023 - targeted invitations & supported completion).

Findings from this analysis will be disseminated by the Patient and Public Involvement group and the research team to relevant stakeholders (midlife women in Brighton and Hove, particularly those in underserved areas and groups, clinicians, commissioners of care, academics) via a scheduled public engagement event (‘Valuing the voices of women in coastal communities’), presentations at conferences, and in a peer-reviewed journal manuscript.

## Data Availability statement

The ‘Health Counts’ data are stored securely and analysed by health researchers at the University of Brighton and Brighton and Sussex Medical School, and by Brighton and Hove City Council’s Public Health Intelligence team. The joint Data Controllers are the University of Brighton and Brighton & Hove City Council. The ‘Health Counts’ dataset is embargoed for a period to be determined by the Editorial Board and cannot be shared with any external party until the embargo period is lifted at which point, access to the dataset will be governed by the Editorial Board procedures.

## Notes

### Competing Interest Statement

The authors have declared no competing interest.

### Funding Statement

Health Counts 2024 was funded by Brighton and Hove City Council Public Health Department. This analysis was funded by a PhD fellowship granted by the University Hospitals Sussex NHS Foundation Trust and Brighton and Sussex Medical School.

### Author Declarations

Ethical approval for the Health Counts survey was obtained from the NHS Health Research Authority (HRA: Ref: 23/LO/0825) and the London Bromley Research Ethics Committee (Ref: 23/LO/0825). Ethical approval for the community enrolment and supported completion pathway was granted by the University of Brighton Cross-School Research Ethics Committee C (2023 to 12553).

